# Diagnostic accuracy of the 4AT for delirium detection: systematic review and meta-analysis

**DOI:** 10.1101/2020.06.11.20128280

**Authors:** Zoë Tieges, Alasdair M. J. MacLullich, Atul Anand, Claire Brookes, Marica Cassarino, Margaret O’Connor, Damien Ryan, Thomas Saller, Rakesh C. Arora, Yue Chang, Kathryn Agarwal, George Taffet, Terence Quinn, Susan. D. Shenkin, Rose Galvin

## Abstract

**Objective:** Detection of delirium in hospitalised older adults is recommended in national and international guidelines. The 4 ‘A’s Test (4AT) is a short (<2 min) instrument for delirium detection that is used internationally as a standard tool in clinical practice. We performed a systematic review and meta-analysis of diagnostic test accuracy of the 4AT for delirium detection.

**Methods:** We searched MEDLINE, EMBASE, PsycINFO, CINAHL, clinicaltrials.gov and the Cochrane Central Register of Controlled Trials, from 2011 (year of 4AT release on the website www.the4AT.com) until 21 December 2019. Inclusion criteria were: older adults (≥ 65y); diagnostic accuracy study of the 4AT index test when compared to delirium reference standard (standard diagnostic criteria or validated tool). Methodological quality was assessed using the Quality Assessment of Diagnostic Accuracy Studies-2 tool. Pooled estimates of sensitivity and specificity were generated from a bivariate random effects model.

**Results:** 17 studies (3702 observations) were included. Settings were acute medicine, surgery, a care home, and the emergency department. Three studies assessed performance of the 4AT in stroke. The overall prevalence of delirium was 24.2% (95% CI 17.8-32.1%; range 10.5-61.9%). The pooled sensitivity was 0.88 (95% CI 0.80-0.93) and the pooled specificity was 0.88 (95% CI 0.82-0.92). Excluding the stroke studies, the pooled sensitivity was 0.86 (95% CI 0.77-0.92) and the pooled specificity was 0.89 (95% CI 0.83-0.93). The methodological quality of studies varied but was moderate to good overall.

**Conclusions:** The 4AT shows good diagnostic test accuracy for delirium in the 17 available studies. These findings support its use in routine clinical practice in delirium detection.

PROSPERO Registration number CRD42019133702.

**Key points:**

- The 4AT is a short delirium assessment tool that is widely used internationally in clinical practice.
- This systematic review and meta-analysis of diagnostic accuracy studies of the 4AT included 3702 observations in 17 studies from nine countries.
- Studies recruited from a range of settings including the Emergency Department, and medical, stroke, and surgical wards.
- The 4AT had a pooled sensitivity of 0.88 and pooled specificity of 0.88.
- The methodological quality of studies varied but was moderate to good overall.

## INTRODUCTION

Delirium is a serious acute neuropsychiatric disorder of consciousness, attention and cognition triggered by general medical conditions, drugs, surgery, or a combination of causes. It manifests through acute and fluctuating cognitive, psychomotor and perceptual disturbances which develop over hours to days. Delirium is common in hospitalised older adults, with a recent meta-analysis of 33 studies of medical inpatients finding an overall delirium occurrence of 23% (95% CI 19-26%) [1]. It is also common in surgical patients, in care homes and palliative care settings [2]. Delirium is associated with significant adverse outcomes including functional decline and mortality, and patient and carer distress [3, 4].

Detection of delirium at the earliest possible time point is important for several reasons, including prompting the search for acute triggers, gaining access to recommended treatment pathways, in managing delirium-associated risks such as falls, in identifying and treating distress, in providing prognostic information, and in communicating the diagnosis to patients and carers. Detection has been recommended in multiple guidelines including the Scottish Intercollegiate Guidelines Network (SIGN) guidelines on delirium [5]. More than 30 delirium assessment tools exist, though these vary considerably in purpose and clinical applicability [6, 7]. Categories of tools include: those intended for episodic use at first presentation or at other points when delirium is suspected; regular use (that is, daily or more frequently) in monitoring for new onset delirium in inpatients; ‘ultra-brief’ screening tools; intensive care unit tools; measurement of delirium severity; informant-based; and detailed phenomenological assessment.

The 4 ‘A’s Test or 4AT was developed as a short delirium assessment tool intended for clinical use in general settings at first presentation and when delirium is suspected. It was initially published on a dedicated website in 2011 [8]. It consists of four items: an item assessing level of alertness, a test of orientation (the Abbreviated Mental Test–4, comprising 4 orientation questions), a test of attention (Months Backward test); and an item ascertaining acute change or fluctuating course [Appendix 1]. The first diagnostic test accuracy study in general settings was published in 2014 [9]. Since publication 4AT performance has been evaluated in multiple studies [10]. The 4AT has become a standard tool in clinical practice [11, 12] and it is recommended in guidelines and pathways [5, 13]. Two prior systematic reviews of general delirium assessment tools included the 4AT but could only cite the original general validation study because of the date of the reviews [6, 7]. A systematic review of delirium detection in stroke patients published in 2019 included three studies that had evaluated the 4AT post-stroke reporting sensitivities from 0.90 to 1.00 and specificities from 0.65 to 0.86 [14].

Here we report a systematic review and meta-analysis of studies that have evaluated the diagnostic test accuracy of the 4AT for delirium detection in older adults in all available care settings.

## METHODS

The methods and search strategy were documented in advance and published in the PROSPERO database (available at http://www.crd.york.ac.uk/PROSPERO/ with registration number CRD42019133702). The review and meta-analysis was conducted in compliance with the principles in the Cochrane Handbook for Systematic Reviews of Diagnostic Test Accuracy [15], and reported using the Preferred Reporting Items for a Systematic Review and Meta-analysis of Diagnostic Test Accuracy Studies (PRISMA-DTA) guidelines [16].

### Search strategy and selection criteria

An inclusive search strategy was developed with a medical librarian. The validated delirium search syntax produced by the National Institute for Health and Clinical Excellence (NICE) clinical guidance for delirium was used to identify delirium [Appendix 2: search strategy]. The following databases were searched: MEDLINE® (OVID), EMBASE (OVID), PsycINFO (EBSCO), CINAHL (EBSCO), clinicaltrials.gov and the Cochrane Central Register of Controlled trials from 2011 (the year the 4AT was published online) to 21 December 2019. We conducted forward citation searches of included articles and checked reference lists of included articles for further articles of potential relevance. We contacted delirium experts from international delirium-focused organisations to identify relevant published or unpublished data and searched relevant conference proceedings.

Studies were included if they met the following criteria: (1) age ≥ 65; (2) examined the diagnostic accuracy of the 4AT for detection of delirium; (3) reference standard assessment of delirium made using standardised diagnostic criteria or a validated tool; and (4) cross-sectional, retrospective or prospective cohort design. If identified studies included adults both younger and older than the threshold age, the study authors were contacted to enquire about the possibility to access data on the older adults only. Studies in patients with delirium tremens were excluded.

### Data extraction

Titles and abstracts were independently screened for inclusion by individuals in pairs of review authors (C.B. and R.G., and Z.T. and A.A.). Full-text screens were carried out independently by two review authors (Z.T. and A.M.). The reviewer pairs performed data extraction independently, resolving disagreement by discussion, or by involving a fifth review author (S.S.) where necessary.

Data were extracted on: type of study; setting; study population; patient demographics; prevalence of delirium; co-morbid illness or illness severity if reported, details of 4AT administration (timing, assessors etc.) and the reference standard; statistics used including adjustments made; and study conclusions. Test accuracy data were extracted to a two-by-two table (number of true positives, false positives, true negatives and false negatives for the 4AT). Study authors were contacted for further information on index and reference test results if insufficient data were provided to perform statistical analyses.

### Risk of bias assessment

Studies were assessed for methodological quality by two independent review authors (R.G. and Z.T.) using the Quality Assessment of Diagnostic Accuracy Studies (QUADAS-2) tool. Narrative summaries were generated describing risk of bias (high, low, or unclear) and concerns regarding applicability. As part of a tailoring phase of the QUADAS-2 tool, the item on the threshold used was omitted because the design of the 4AT pre-specifies the threshold to be used for delirium detection (cut-off ≥ 4). For the item on the appropriate interval between index test and reference standard, the interval was set to a maximum of three hours [Appendix 3: Assessment of methodological quality with the QUADAS-2 tool].

### Statistical analysis

Meta-analyses were completed using Diagnostic Test Accuracy Meta-Analysis software, version 1.21 (https://crsu.shinyapps.io/dta_ma/) [17]. Pooled estimates of delirium prevalence were calculated using random effects models (*meta* package in R [18]). The primary outcome of interest was the identification of delirium (presented as a dichotomous yes/no variable) by a reference standard (i.e. Diagnostic and Statistical Manual of Mental Disorders (DSM)) or a validated diagnostic tool such as the Confusion Assessment Method (CAM) [19]. Summary estimates of sensitivity and specificity with 95% confidence intervals (CI) were calculated using a bivariate random effects model. Receiver operating characteristic (ROC) curves were used to plot summary estimates of sensitivity and specificity.

A sensitivity analysis was performed including only those studies which were deemed to have an overall low risk of bias (that is, high study quality). Pre-planned subgroup analyses were also conducted to investigate clinical heterogeneity across studies: (i) excluding studies in patients with stroke, because of the potential influence of aphasia on the test, [20, 21], to assess test accuracy of the 4AT in non-stroke populations, and (ii) analysing separately for studies using (a) a clinical reference standard (e.g. DSM) or (b) a validated assessment tool (e.g. the Confusion Assessment Method (CAM)). A post-hoc subgroup analysis was conducted to compare diagnostic accuracy of the English 4AT versus the translated versions.

## RESULTS

### Study identification

We identified 853 records from our initial search and 3 records from conference abstracts (Figure 1). A total of 780 records remained after initial deduplication. Following title and abstract screening, 21 records had full-text review and 16 articles were included reporting 17 different studies [9, 10, 22-35]. The main reason for exclusion of articles was that studies were not designed as a diagnostic accuracy study of the 4AT and/or did not include data that allowed derivation of diagnostic test accuracy data. One conference proceeding reported two separate studies [25]. Three authors provided study data on subgroups of older patients [23, 27, 30, 36].

**Figure 1.**
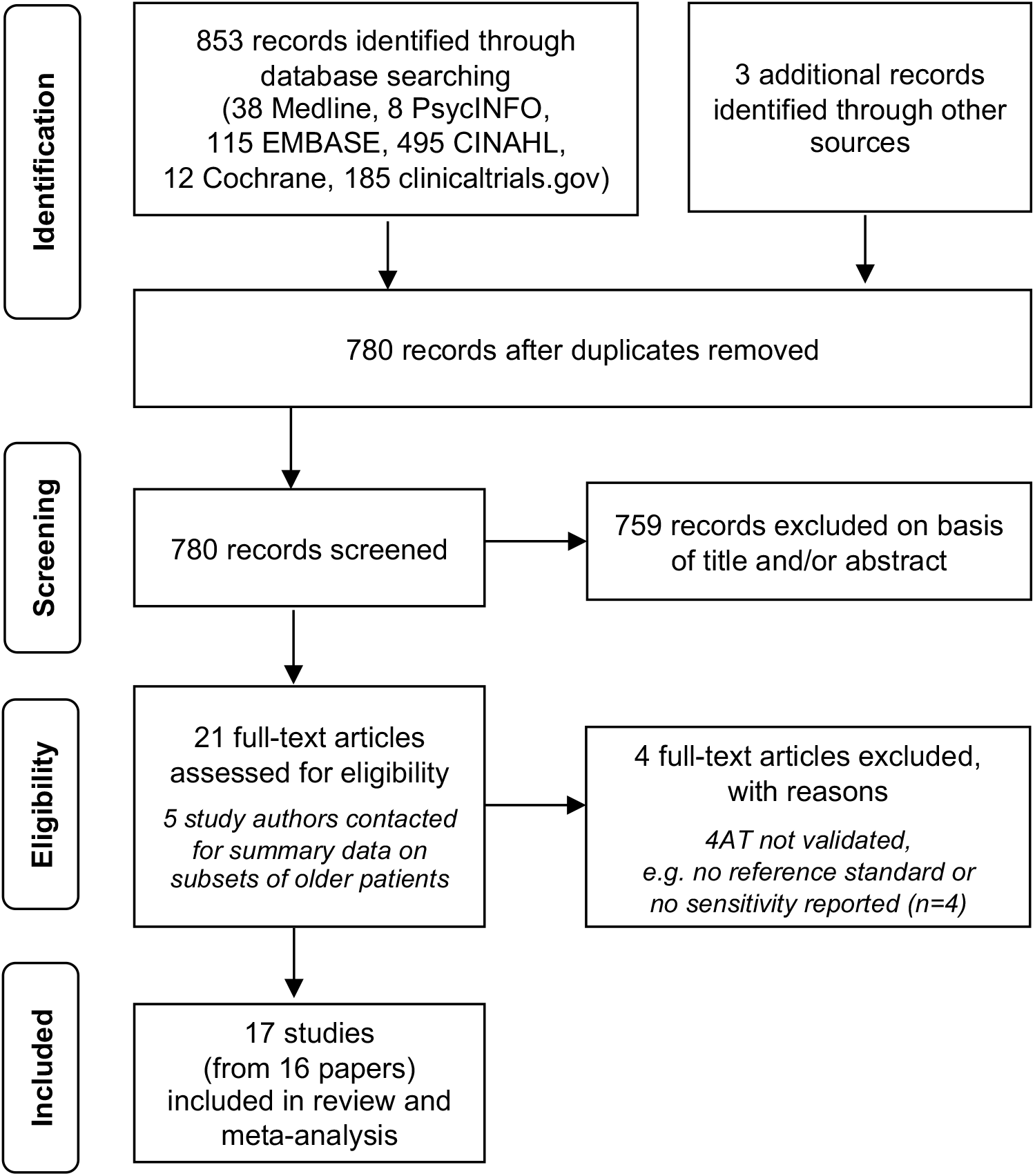
PRISMA flow chart diagram for the search and study selection process. PRISMA: *P*referred *R*eporting *I*tems for *S*ystematic Reviews and *M*eta-*A*nalyses.

### Study characteristics

A summary of the characteristics of the included studies is provided in Table 1. The number of study participants ranged between 49 [26] and 785 [10]. The prevalence of delirium across the studies was 24.2% (95% CI 17.8-32.1%), varying between 10.5% [36] to 61.9% [28]. Eight studies validated a translated version of the 4AT in Italian, Persian, Thai, Russian, French, Norwegian and German [9, 22, 24, 26, 29-31, 36]. Two studies used a modified 4AT where the months of the year backwards test was replaced by the days of the week backwards to assess inattention [25, 32]; this modification does not affect the threshold scoring for delirium versus no delirium in the tool. Studies were conducted in inpatient general medical or geriatric medical wards, acute stroke units, emergency departments and post-operative care units, and nursing homes, in eleven countries. In one study in Australia 39% of participants were non-English speakers and required an interpreter during the assessment [28].

**Table 1:**
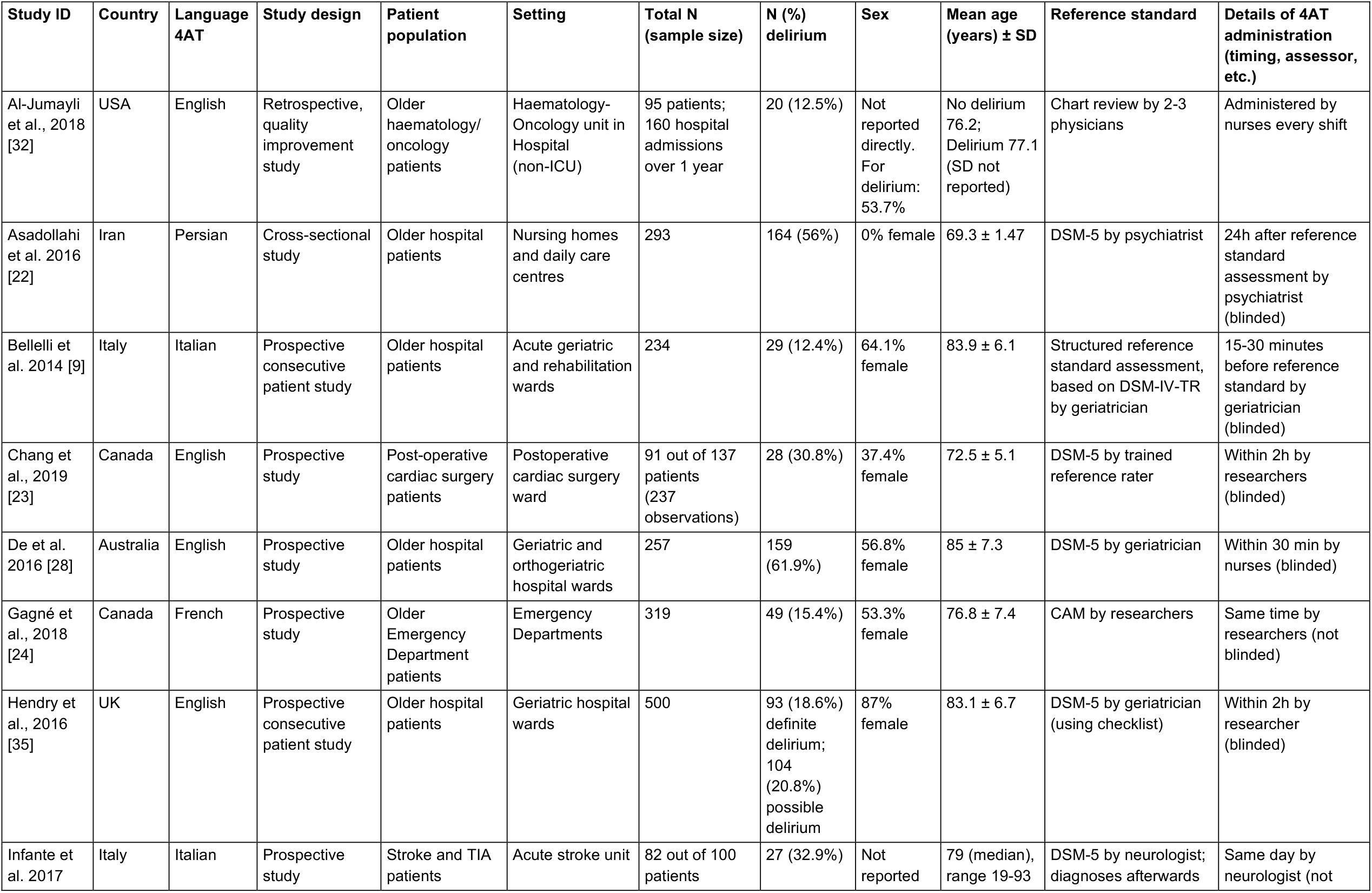
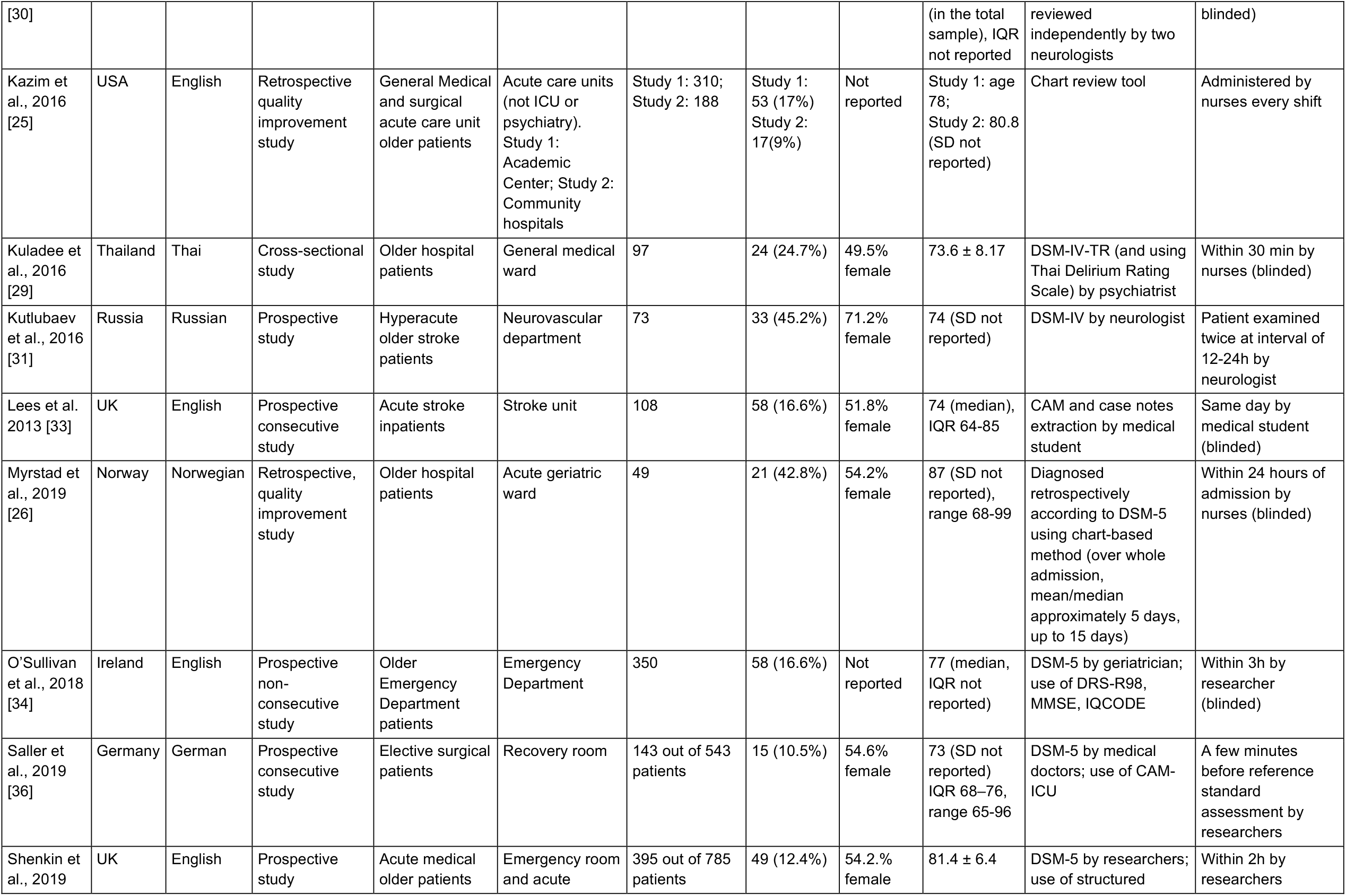
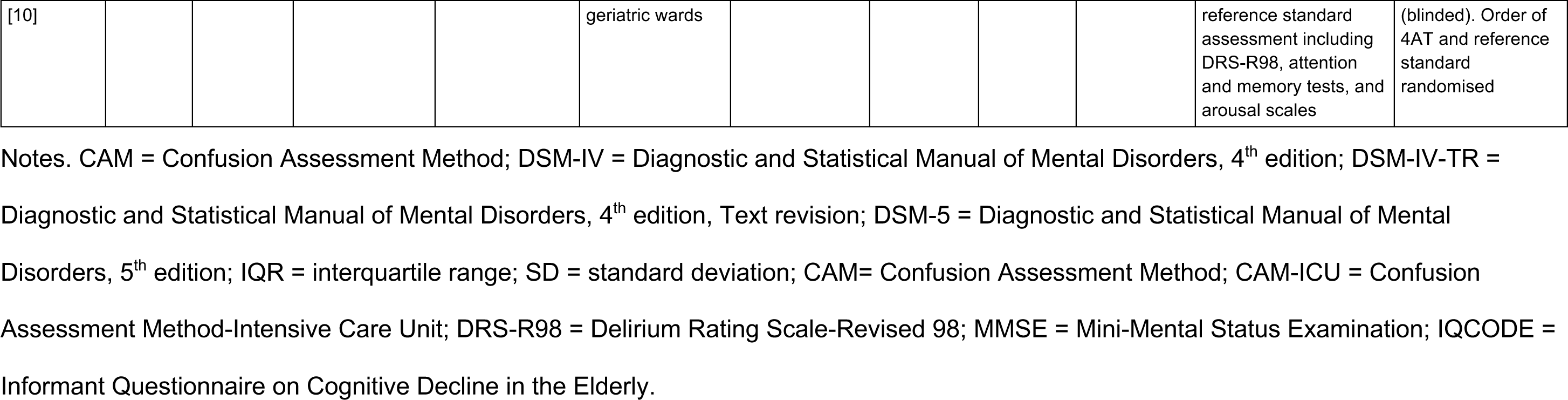
Characteristics of included studies

| Study ID | Country | Language 4AT | Study design | Patient population | Setting | Total N (sample size) | N (%) delirium | Sex | Mean age (years) ± SD | Reference standard | Details of 4AT administration (timing, assessor, etc.) |
| --- | --- | --- | --- | --- | --- | --- | --- | --- | --- | --- | --- |
| Al-Jumayli et al., 2018 [32] | USA | English | Retrospective, quality improvement study | Older haematology/ oncology patients | Haematology- Oncology unit in Hospital (non-ICU) | 95 patients; 160 hospital admissions over 1 year | 20 (12.5%) | Not reported directly. For delirium: 53.7% | No delirium 76.2; Delirium 77.1 (SD not reported) | Chart review by 2-3 physicians | Administered by nurses every shift |
| Asadollahi et al. 2016 [22] | Iran | Persian | Cross-sectional study | Older hospital patients | Nursing homes and daily care centres | 293 | 164 (56%) | 0% female | 69.3 ± 1.47 | DSM-5 by psychiatrist | 24h after reference standard assessment by psychiatrist (blinded) |
| Bellelli et al. 2014 [9] | Italy | Italian | Prospective consecutive patient study | Older hospital patients | Acute geriatric and rehabilitation wards | 234 | 29 (12.4%) | 64.1% female | 83.9 ± 6.1 | Structured reference standard assessment, based on DSM-IV-TR by geriatrician | 15-30 minutes before reference standard by geriatrician (blinded) |
| Chang et al., 2019 [23] | Canada | English | Prospective study | Post-operative cardiac surgery patients | Postoperative cardiac surgery ward | 91 out of 137 patients (237 observations) | 28 (30.8%) | 37.4% female | 72.5 ± 5.1 | DSM-5 by trained reference rater | Within 2h by researchers (blinded) |
| De et al. 2016 [28] | Australia | English | Prospective study | Older hospital patients | Geriatric and orthogeriatric hospital wards | 257 | 159 (61.9%) | 56.8% female | 85 ± 7.3 | DSM-5 by geriatrician | Within 30 min by nurses (blinded) |
| Gagné et al., 2018 [24] | Canada | French | Prospective study | Older Emergency Department patients | Emergency Departments | 319 | 49 (15.4%) | 53.3% female | 76.8 ± 7.4 | CAM by researchers | Same time by researchers (not blinded) |
| Hendry et al., 2016 [35] | UK | English | Prospective consecutive patient study | Older hospital patients | Geriatric hospital wards | 500 | 93 (18.6%) definite delirium; 104 (20.8%) possible delirium | 87% female | 83.1 ± 6.7 | DSM-5 by geriatrician (using checklist) | Within 2h by researcher (blinded) |
| Infante et al. 2017 | Italy | Italian | Prospective study | Stroke and TIA patients | Acute stroke unit | 82 out of 100 patients | 27 (32.9%) | Not reported | 79 (median), range 19-93 | DSM-5 by neurologist; diagnoses afterwards | Same day by neurologist (not |
| [30] |  |  |  |  |  |  |  |  | (in the total sample), IQR not reported | reviewed independently by two neurologists | blinded) |
| Kazim et al., 2016 [25] | USA | English | Retrospective quality improvement study | General Medical and surgical acute care unit older patients | Acute care units (not ICU or psychiatry). Study 1: Academic Center; Study 2: Community hospitals | Study 1: 310; Study 2: 188 | Study 1: 53 (17%)<br>Study 2: 17(9%) | Not reported | Study 1: age 78;<br>Study 2: 80.8 (SD not reported) | Chart review tool | Administered by nurses every shift |
| Kuladee et al., 2016 [29] | Thailand | Thai | Cross-sectional study | Older hospital patients | General medical ward | 97 | 24 (24.7%) | 49.5% female | 73.6 ± 8.17 | DSM-IV-TR (and using Thai Delirium Rating Scale) by psychiatrist | Within 30 min by nurses (blinded) |
| Kutlubayev et al., 2016 [31] | Russia | Russian | Prospective study | Hyperacute older stroke patients | Neurovascular department | 73 | 33 (45.2%) | 71.2% female | 74 (SD not reported) | DSM-IV by neurologist | Patient examined twice at interval of 12-24h by neurologist |
| Lees et al. 2013 [33] | UK | English | Prospective consecutive study | Acute stroke inpatients | Stroke unit | 108 | 58 (16.6%) | 51.8% female | 74 (median), IQR 64-85 | CAM and case notes extraction by medical student | Same day by medical student (blinded) |
| Myrstad et al., 2019 [26] | Norway | Norwegian | Retrospective, quality improvement study | Older hospital patients | Acute geriatric ward | 49 | 21 (42.8%) | 54.2% female | 87 (SD not reported), range 68-99 | Diagnosed retrospectively according to DSM-5 using chart-based method (over whole admission, mean/median approximately 5 days, up to 15 days) | Within 24 hours of admission by nurses (blinded) |
| O'Sullivan et al., 2018 [34] | Ireland | English | Prospective non-consecutive study | Older Emergency Department patients | Emergency Department | 350 | 58 (16.6%) | Not reported | 77 (median, IQR not reported) | DSM-5 by geriatrician; use of DRS-R98, MMSE, IQCODE | Within 3h by researcher (blinded) |
| Saller et al., 2019 [36] | Germany | German | Prospective consecutive study | Elective surgical patients | Recovery room | 143 out of 543 patients | 15 (10.5%) | 54.6% female | 73 (SD not reported) IQR 68–76, range 65-96 | DSM-5 by medical doctors; use of CAM-ICU | A few minutes before reference standard assessment by researchers |
| Shenkin et al., 2019 | UK | English | Prospective study | Acute medical older patients | Emergency room and acute | 395 out of 785 patients | 49 (12.4%) | 54.2.% female | 81.4 ± 6.4 | DSM-5 by researchers; use of structured | Within 2h by researchers |
| [10] |  |  |  |  | geriatric wards |  |  |  |  | reference standard<br>assessment including<br>DRS-R98, attention<br>and memory tests, and<br>arousal scales | (blinded). Order of<br>4AT and reference<br>standard<br>randomised |

Notes. CAM = Confusion Assessment Method; DSM-IV = Diagnostic and Statistical Manual of Mental Disorders, 4<sup>th</sup> edition; DSM-IV-TR = Diagnostic and Statistical Manual of Mental Disorders, 4<sup>th</sup> edition, Text revision; DSM-5 = Diagnostic and Statistical Manual of Mental Disorders, 5<sup>th</sup> edition; IQR = interquartile range; SD = standard deviation; CAM= Confusion Assessment Method; CAM-ICU = Confusion Assessment Method-Intensive Care Unit; DRS-R98 = Delirium Rating Scale-Revised 98; MMSE = Mini-Mental Status Examination; IQCODE = Informant Questionnaire on Cognitive Decline in the Elderly.

### Study quality

The methodological quality of studies varied but was moderate to good overall. Potential for bias in studies was generally low, but where present was due to the selection of participants (excluding patients unable to give consent or those with dementia, n=2), the timing between the reference standard assessment and the 4AT (not reported (n=6) or exceeding the maximum interval of 3 hours (n=2)), and the blinding of assessments (unblinded raters (n=2) or blinding status unclear (n=3)) (Table 1 and Figure 2). Seven papers were of higher concern (rated high or unclear risk of bias across three areas). Nine studies were considered low risk overall.

**Figure 2.**
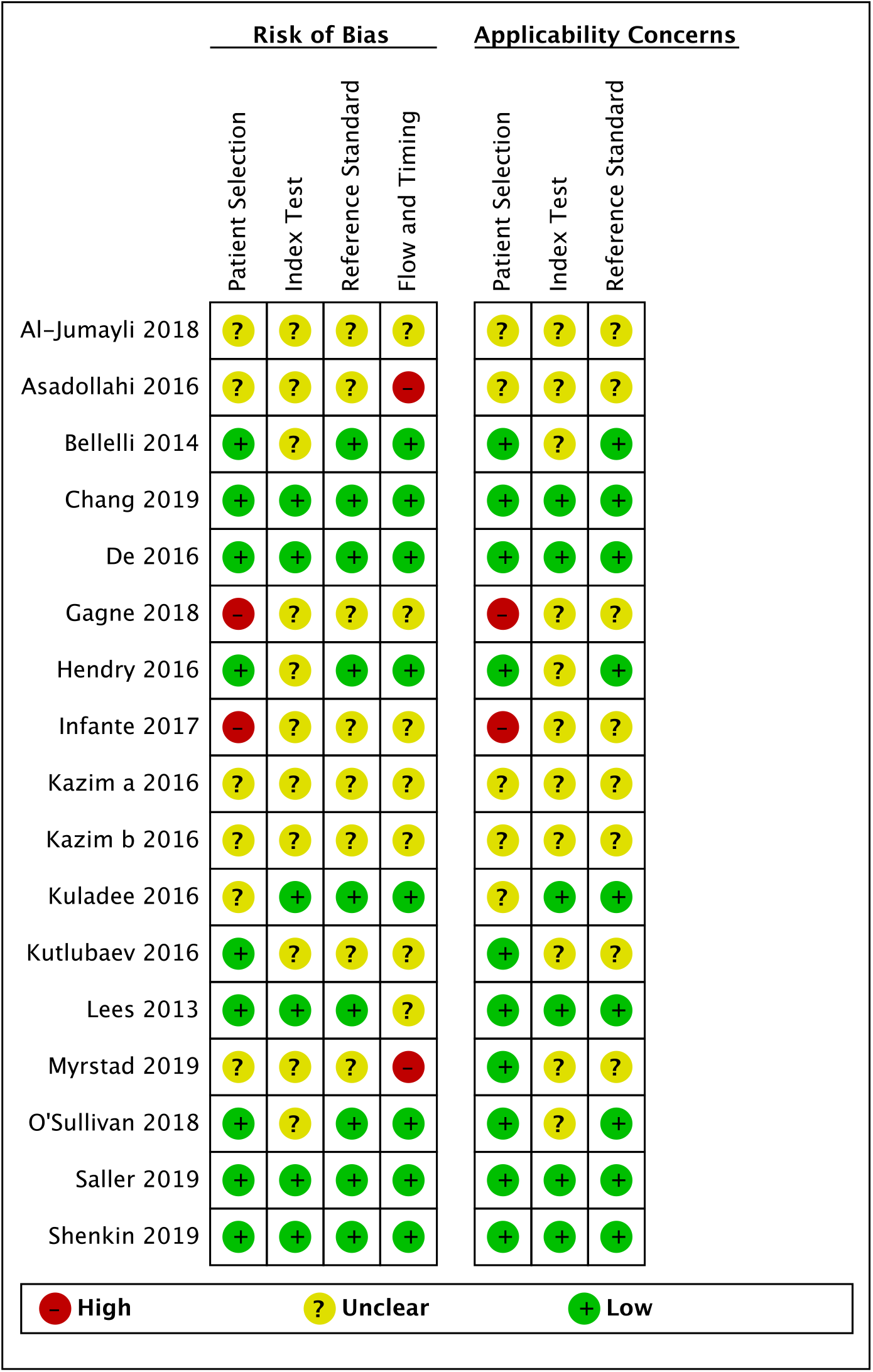
Risk of bias and applicability concerns summary (see Appendix 3 for QUADAS-2 assessment criteria).

Myrstad et al. [26] used a reference standard based on the whole length of stay (median 5 days) whereas the 4AT was performed in the first 24 hours of the admission; this could have led to a reduced sensitivity of the 4AT as some delirium arises after the first 24 hours. Hendry et al. [35] administered the 4AT as part of a larger cognitive test battery, therefore the index rater had knowledge of the participant’s mental status beyond that elicited by the 4AT assessment that could have affected 4AT scoring. Gagné et al. [24] repeatedly administered the 4AT and the combined results were incorporated in the reported sensitivity and specificity. Asadollahi et al. [22] administered the 4AT only to those patients who had delirium according to DSM-5 criteria.

### Diagnostic test accuracy

All 17 studies were included in the meta-analysis. The 4AT had a pooled sensitivity for detecting delirium of 0.88 (95% CI 0.80-0.93) and a pooled specificity of 0.88 (95% CI 0.82-0.92), indicating good diagnostic accuracy of the 4AT as a tool to identify individuals at high risk of delirium (Table 2 and Figure 3). These estimates were broadly consistent across studies with the exception of two studies reporting lower sensitivities (both studies had a high or unclear risk of bias) [22, 26]. A sensitivity analysis including 9 studies with overall low risk of bias resulted in comparable summary estimates of sensitivity (0.87, 95% CI 0.84-0.90) and specificity (0.88, 95% CI 0.81-0.93) [Appendix 5].

**Table 2.** Summary estimates of sensitivity and specificity.

| Application<br>of 4AT | No. of studies<br>(observations) | Sensitivity<br>(95% CI) | Specificity<br>(95% CI) |
| --- | --- | --- | --- |
| All studies | 17 (3702) | 0.88<br>(0.80-0.93) | 0.88<br>(0.82-0.92) |
| Sensitivity<br>analysis (low<br>risk of bias) | 9 (2252) | 0.87<br>(0.84-0.90) | 0.88<br>(0.81-0.93) |
| Subgroup<br>analysis<br>(excluding<br>stroke) | 14 (3440) | 0.86<br>(0.77-0.92) | 0.89<br>(0.83-0.93) |

**Figure 3.**
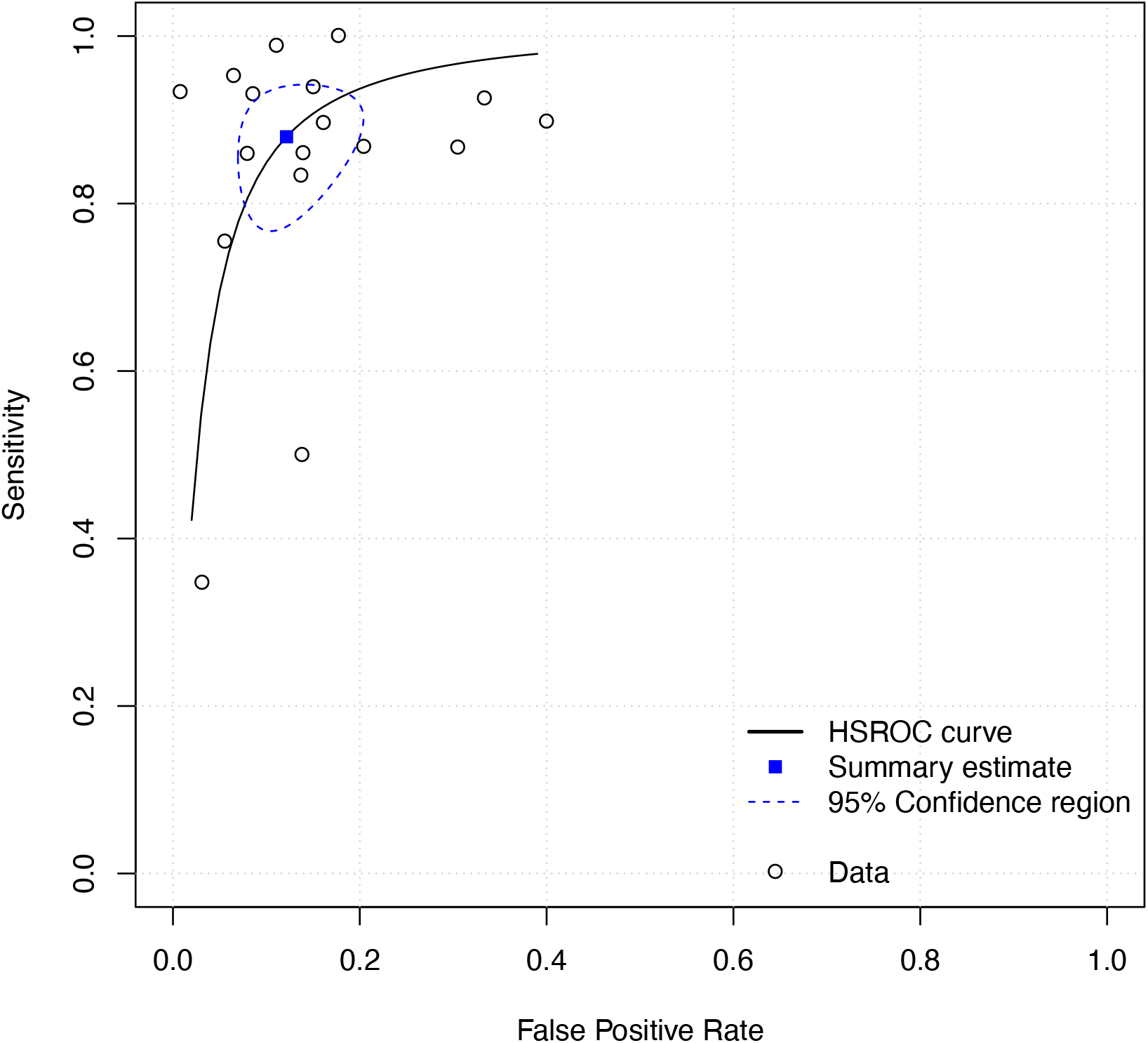
Hierarchical Summary Receiver Operating Characteristic (HSROC) curve of the 4AT for identifying individuals with delirium.

The planned subgroup analysis excluding three studies in stroke patients resulted in similar summary estimates of sensitivity (0.86; 95% CI 0.77-0.92) and specificity (0.89; 95% CI 0.83-0.93), suggesting robust results across populations (results of the other subgroup analyses are presented in Appendix 4]. Three studies reported findings in subsets of patients with known dementia, with sensitivities of 0.94, 0.86, and 0.92 and specificities of 0.65, 0.71, and 0.79, in the Bellelli et al. [9], De et al. [28], and O’Sullivan et al. [34] studies, respectively.

## DISCUSSION

### Statement of principal findings

This systematic review identified 17 studies involving 3702 observations evaluating the diagnostic test accuracy of the 4AT for detection of delirium in older patients (≥ 65y) across eleven countries, a variety of care settings and in multiple languages. The prevalence of delirium was 24.2% (N=945), ranging from 10.5%-61.9%. Pooled sensitivity and specificity were 0.88 and 0.88, respectively, indicating good accuracy. Notably, the sensitivity and specificity were balanced. Similar estimates were demonstrated when subgroup analyses were performed based on study quality and population type.

### Results in the context of the current literature

Delirium detection remains a major challenge, with recent studies continuing to show underdetection [37]. An important factor in improving detection is the availability of validated assessment tools usable in clinical practice. The 4AT now has a substantial evidence base supporting its validity as a delirium assessment tool. Coupled with this is also emerging evidence of implementation of the 4AT scale in routine clinical practice, for example in data from the National Hip Fracture Database which assesses the clinical care of >95% hip fracture patients in England, Wales and Northern Ireland. In 2018, 25% of approximately 60,000 4AT assessments (92% of all patients) performed in the 7 postoperative days (the audit period) were positive [11]. Though diagnostic accuracy data were not collected, these data suggest that the 4AT may be detecting the expected level of delirium.

There are many other tools in the literature; however those with profiles of intended use similar to the 4AT with more than two published diagnostic accuracy studies are the CAM, the 3D-CAM, and the brief CAM (bCAM) [6, 35, 38-44]. The CAM was first published in 1990 and is a widely used tool in research and clinical practice. There are 23 published CAM diagnostic test accuracy studies involving a total of 2629 patients [10, 38], with sensitivities of 0.09-1.0 and specificities of 0.84-1.0 reported. There is limited published information on its performance in routine clinical care. One recent large study found a sensitivity of 0.27 [39] though the CAM was scored without the recommended preceding interview and cognitive testing. Alternative tools include the 3D-CAM, a 20-item variant of the CAM that takes 2-5 minutes to complete (median 3 minutes) [40], and the bCAM, a 2 minute, 4-item variant of the CAM designed and validated for use in the emergency department [41]. Both of these tools show generally good performance in published diagnostic test accuracy studies, with reported 3D-CAM sensitivities of 0.85-1.0 and specificities of 0.88-0.97 [40, 45-47], and reported bCAM sensitivities of 0.65-0.84 and specificities of 0.87-0.97 [35, 41-44]. To our knowledge there are currently no published clinical implementation data for these tools.

Our review provides evidence that the 4AT has good diagnostic test accuracy for identification of delirium, with a body of validation data comparable to the CAM. The 4AT has some advantages over the CAM and 3D-CAM, being shorter and simpler, and not requiring special training. Notably, the 4AT had a higher sensitivity than the CAM, though with similar specificity, in a recent STARD-compliant randomised controlled trial [10]. As with other delirium tools, studies on clinical implementation of the 4AT are relatively lacking. These kinds of studies might expose training needs or other challenges in implementation such as lower sensitivity when used in routine practice. Additionally, the 4AT lacks diagnostic accuracy data in palliative care settings and has limited data in the community. The number of studies examining its performance in patients with known dementia is relatively small; the three studies presented in this review found lower specificity in delirium superimposed on dementia [9, 28, 34].

### Strengths and weaknesses of the study

This is the first meta-analysis of 4AT diagnostic test accuracy studies. Our findings were broadly consistent across different care settings and languages. We published the protocol in advance, and we used systematic and robust methods including using a comprehensive search strategy, and independent reviewers to identify, select, appraise and synthesise relevant studies. The selected studies originated from nine countries, and eight were conducted with a translated version of the tool. Thus, the findings of the review suggest good generalisability. The methodological quality of the studies was moderate to good overall, despite some uncertainty in relation to the conduct of the 4AT in four studies. The two studies showing low sensitivities both had high risk of bias. Due to the data of the studies included in this review, it was not possible to perform sensitivity analyses to determine the impact of time interval between tests and this should be the subject of further studies. Also, the Cochrane guidelines recommend the use of a single reference standard in order to prevent bias or ambiguity, but we included studies using either DSM-IV, DSM-5 or CAM as reference standard to maximise comprehensiveness.

### Areas for further research

Methodological deficiencies related to the timings of the reference standard and 4AT identified in this review, as well as lack of adherence to the STARD guidelines, should be addressed in future validation studies. Studies evaluating the 4AT in other settings and in patients with dementia, preferably taking into account the severity of dementia, are required. Clinical implementation studies evaluating 4AT performance including completion rates as well as diagnostic accuracy in routine clinical practice are also needed.

## Conclusion

This meta-analysis quantifies the diagnostic accuracy of the 4AT. The psychometric performance is good and coupled with its simplicity and brevity, the present findings support use of the 4AT in routine clinical practice.

## Data Availability

All relevant data are provided within the paper.

## List of abbreviations

3D-CAM: 3-Minute Diagnostic Assessment for Delirium using the Confusion Assessment Method algorithm
4AT: 4 ‘A’s Test
bCAM: brief Confusion Assessment Method
CAM-ICU: Confusion Assessment Method-Intensive Care Unit
CAM: Confusion Assessment Method
DRS-R98: Delirium Rating Scale-Revised 98
DSM: Diagnostic and Statistical Manual of Mental Disorders
IQCODE: Informant Questionnaire on Cognitive Decline in the Elderly
MMSE: Mini-Mental Status Examination
NICE: National Institute for Health and Clinical Excellence
PRISMA-DTA: Preferred Reporting Items for a Systematic Review and Meta-analysis of Diagnostic Test Accuracy Studies
QUADAS-2: Quality Assessment of Diagnostic Accuracy Studies-Version 2
SIGN: Scottish Intercollegiate Guidelines Network
STARD: Standards for Reporting Diagnostic accuracy studies

## Appendix 1. The 4AT

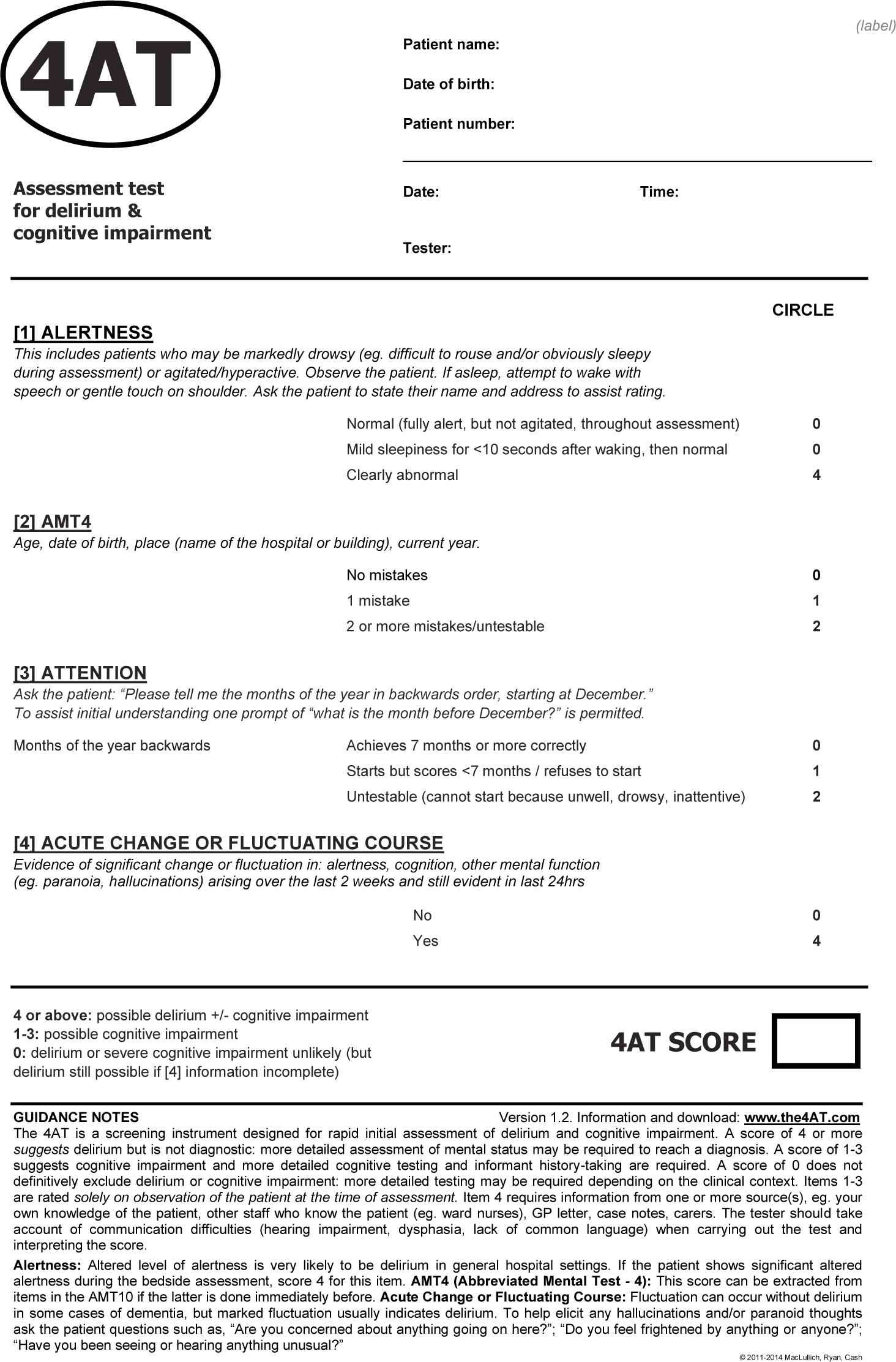

Note: The 4AT can be downloaded from www.the4at.com in different languages.

## Appendix 2: Search strategy

MEDLINE Search terms

“4 A’s test”.mp.

2. “four A’s Test”.mp.

3. “4 ‘A’s Test”.mp.

4. “4 A-T”.mp.

5. “four A-T”.mp.

6. “4-A Test”.mp.

7. “4 A’s scale”.mp.

8. “four ‘A’s test”.mp.

9. ‘4AT’.mp.

10. 1 or 2 or 3 or 4 or 5 or 6 or 7 or 8 or 9

11. limit 10 to yr=“2011 -Current”

12. deliri$.ti,ab.

13. (acute adj2 (confusion$ or “brain syndrome” or “brain failure” or “psycho-organic syndrome” or “organic psychosyndrome”)).mp.

14. (terminal$ adj restless$).mp.

15. toxic confus$.mp.

16. delirium/

17. confusion/

18. 12 or 13 or 14 or 15 or 16 or 17

19. 11 and 18

Note: The delirium search strategy was taken from: https://www.nice.org.uk/guidance/cg103/documents/delirium-appendix-c-search-strategies2. The strategies for other databases are available on request.

Conference proceedings from the following professional societies were searched: Scottish Delirium Association; European Delirium Association (EDA); American Delirium Society (ADS); and Australasian Delirium Association (ADA). Members of the EDA, ADS and ADA were also contacted via email and twitter to identify relevant published or unpublished data.

## Appendix 3.

**Supplementary Table S1.** Assessment of methodological quality with the QUADAS-2 tool.

| **Domain** | **Patient selection** | **Index test** | **Reference standard** | **Flow and timing** |
| --- | --- | --- | --- | --- |
| Description | Describe methods of patient selection: describe included patients (prior testing, presentation, intended use of index test and setting). | Describe the index test and how it was conducted and interpreted. | Describe the reference standard and how it was conducted and interpreted. | Describe any patients who did not receive the index test(s) and/or reference standard or who were excluded from the 2 x 2 table (refer to flow diagram): describe the time interval and any interventions between index test(s) and reference standard. |
| Signalling questions<br>(yes, no, unclear) | Was a consecutive or random sample of patients enrolled? | Were the index test results interpreted without knowledge of the results of | Is the reference standard likely to correctly classify the target condition? | Was there an appropriate interval between index test(s) and reference |
|  | Was a case-control design avoided?<br>Did the study avoid inappropriate exclusions? | the reference standard?<br>Was the person who administered 4AT not trained/not expert in delirium? | Were the reference standard results interpreted without knowledge of the results of the index test? | standard (3h maximum)?<br>Did all patients receive the same reference standard?<br>Were all patients included in the analysis? |
| Risk of bias | Could the selection of patients have introduced bias? | Could the conduct or interpretation of the index test have introduced bias? | Could the reference standard, its conduct, or its interpretation have introduced bias? | Could the patient flow have introduced bias? |
| Concerns regarding applicability | Are there concerns that the included patients do not match the review question? | Are there concerns that the index test, its conduct, or its interpretation differ from the review question? | Are there concerns that the target condition as defined by the reference standard does not match the review question? | — |

## Appendix 4.

**Supplementary Table S2.** Summary estimates of sensitivity and specificity per subgroup.

| Subgroup | No. of studies<br>(observations) | Sensitivity<br>(95% CI) | Specificity<br>(95% CI) |
| --- | --- | --- | --- |
| Studies using a clinical reference standard (e.g. DSM) | 12 (2640) | 0.84<br>(0.73-0.90) | 0.89<br>(0.82-0.93) |
| Studies using a validated assessment tool | 5 (1062) | 0.95<br>(0.86-0.98) | 0.86<br>(0.75-0.93) |
| Studies using the English 4AT | 9 (2413) | 0.91<br>(0.84-0.94) | 0.88<br>(0.83-0.92) |
| Studies using a translated version of the 4AT | 8 (1289) | 0.83<br>(0.66-0.92) | 0.88<br>(0.76-0.95) |

## Appendix 5.

**Supplementary Figure.**
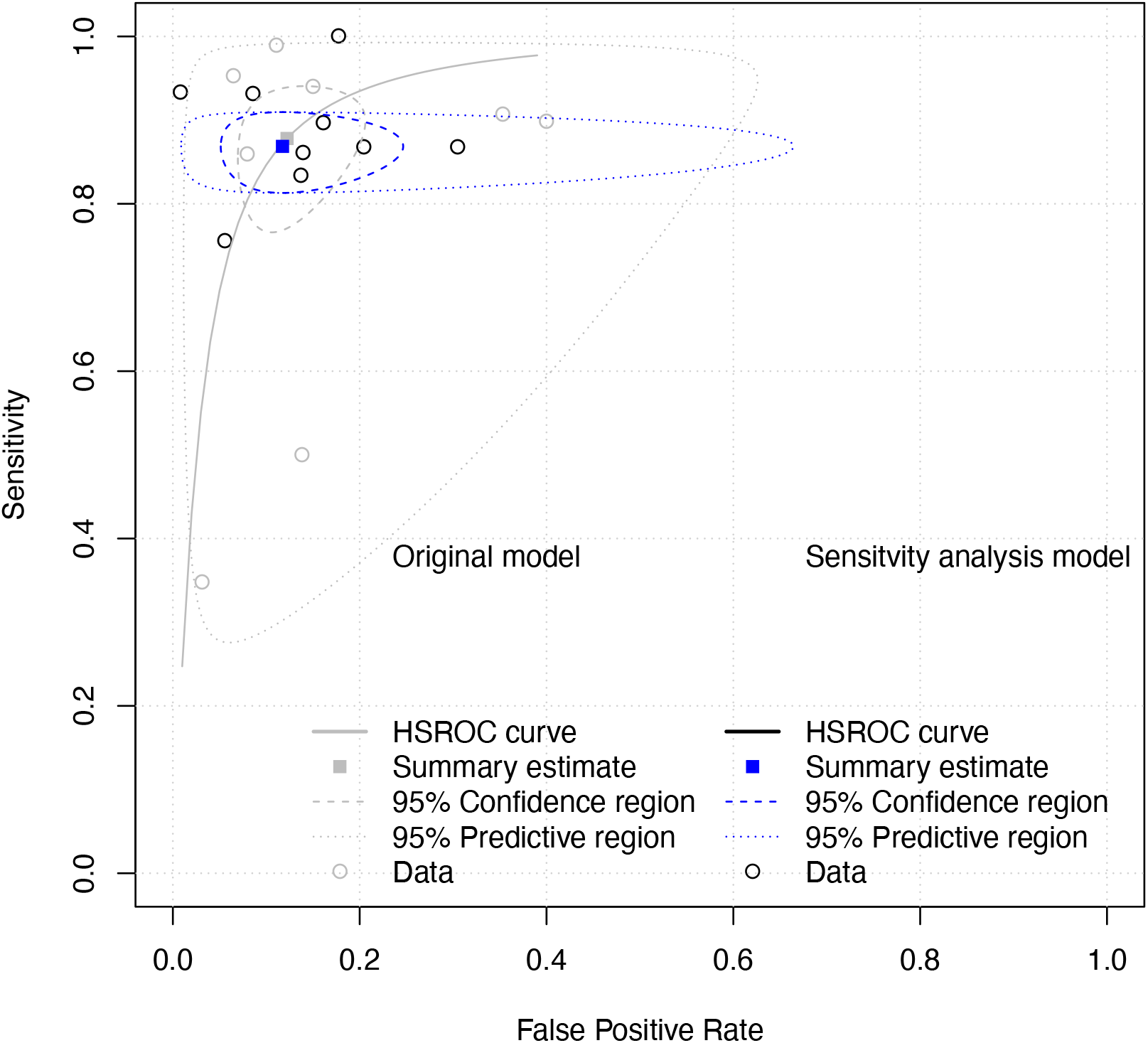
Hierarchical Summary Receiver Operating Characteristic (HSROC) curve of the 4AT for identifying individuals with delirium: results from sensitivity analysis.

The sensitivity analysis included 9 studies rated as low risk of bias. Hierarchical Summary Receiver Operating Characteristic (HSROC) curve of the 4AT for identifying individuals with delirium: the bivariate summary estimates (solid ellipses), with the corresponding 95% prediction ellipses (outer dotted lines) and 95% confidence ellipses (inner dashed lines).

